# Design and methodology of a randomized clinical trial of prolonged daily antibiotic suppression with and without fulguration for uncomplicated recurrent urinary tract infections in women

**DOI:** 10.64898/2026.05.11.26352945

**Authors:** Philippe E. Zimmern, Colby E. Souders, Bonnie C. Prokesch, Kevin C. Lutz, Nicole J. De Nisco

## Abstract

**Objective:** Recurrent urinary tract infections (rUTIs) significantly decrease quality of life and antibiotics are becoming increasingly less effective due to antimicrobial resistance. Alternative effective treatment strategies are urgently needed for rUTIs. Prior studies have indicated that women can experience resolved or improved rUTI following electrofulguration (EF). To further investigate these findings, we report on the design and methodology behind a randomized trial examining two treatment arms: standard prolonged antibiotic treatment with nitrofurantoin (NF) alone or in combination with EF.

**Patients and Methods:** The aim of this randomized trial is to determine, at two institutions, the efficacy of two interventions for rUTI associated with early stages of chronic cystitis (stages 1 and 2): conventional 6 months low-dose (100mg) NF daily antibiotic suppression alone (NF) or conventional NF with EF (EF + NF). The study is also designed to analyze changes in the urinary microbiomes in the two different treatment arms and to determine the durability of clinical outcomes in both treatment arms at 2 years after the end of each intervention.

The primary outcomes will be obtained from 6 to 18 months, as well as 18 – 30 months following completion of the original 6-month intervention. Failure is defined based on UTI symptoms documented by a validated questionnaire with a documented urine culture confirming a bacterial strain at each UTI episode following the end of the 6-month intervention.

**Conclusions:** This randomized trial is designed to examine the efficacy and durability of treating women with rUTIs using the standard of care of NF alone, or an EF procedure with NF.

## Introduction

Urinary tract infections (UTIs) result in considerable morbidity and healthcare expenditures, especially for those who suffer from recurrent UTIs (rUTIs), defined as three or more symptomatic UTIs in a 12-month period or two or more in a six-month period.^1–4^ Recent research has determined the considerable physical and psychological toll of UTIs,^5^ and demonstrated the frequency and impacts of antibiotic resistance and allergy in patients with rUTIs.^6^ Low-dose long-term antibiotic prophylaxis is one of the strategies typically considered for individuals who have frequent UTIs and have not durably responded to several 5-7-day courses of antibiotics.^7^

To investigate the source of reinfection in rUTI sufferers, our group examined the bladder wall during office cystoscopy and observed evidence of chronic inflammatory changes over the trigone and bladder base.^8^ Using a simple outpatient cystoscopic procedure, electrofulguration (EF), or cauterization of these areas of chronic inflammation within the bladder, we have shown symptomatic relief, endoscopic resolution of cystitis lesions, and clinical reduction in rUTIs.^9–11^ Within bladder biopsies obtained in postmenopausal women with rUTI undergoing EF, 16S rRNA fluorescence *in situ* hybridization (FISH) identified resident bacteria in inflamed regions of the bladder wall.^12^ Combined with a more recent study identifying tissue-resident *Escherichia* in these biopsies using genus-specific probes, these findings support bladder reservoirs as a potential source of reinfection.

We recently reported long-term outcomes after EF (median follow-up 11 years) in women with stage 1 and 2 cystitis and a history of uncomplicated rUTI.^13^ These women had a negative upper and lower urinary tract evaluation. Therefore, chronic cystitis represented the only identifiable abnormality associated with UTI recurrences. In this patient cohort, 72% of women were cured, 22% were improved, and 6% did not benefit from the procedure.^13^ Despite our promising results, large gaps in knowledge in the management of women with rUTI indicate the need for definitive Level I evidence comparing the benefits of guideline-driven management relying on long-term (six months) daily antibiotic prophylaxis versus EF of areas of chronic cystitis combined with long-term (six months) daily antibiotic prophylaxis in this population. Thus, our central hypothesis is that those treated with the addition of EF will experience superior reduction in UTI episodes in the short and long-terms compared to those undergoing the standard antibiotic therapy alone.

Here, we describe a multicenter, randomized trial to determine whether EF with six-month antibiotic prophylaxis will reduce UTI episodes over one- and two-year follow-up periods compared to six-month antibiotic prophylaxis alone in women with a well-documented history of uncomplicated rUTI. Additionally, we designed the study to determine changes in the urinary microbiomes following EF with six-month antibiotic prophylaxis compared to six-month antibiotic prophylaxis alone.

## Methods

This trial is being conducted by the University of Texas Southwestern (UTSW) and University of Kansas Medical Center (KUMC), along with a biostatistical coordinating center at UTSW. The work is sponsored by the National Institute of Health (NIH) and the protocol was approved by the Institutional Review Board (IRB) at each of the participating centers. Both interventions (NF pills and EF procedure) are fully funded by the trial at no cost to the participants, and their follow-up visits are compensated to encourage compliance with the study up to two years after the completion of each treatment arm.

### Study Design

This is a two-arm, unmasked, multicenter, randomized clinical trial (**Figure 1**) comparing traditional antibiotic treatment (NF) with and without EF in women with uncomplicated rUTI. There are three primary goals of this study: 1) to compare rUTI episodes for the duration of this study (up to two years) in those treated solely with six months of low dose antibiotic prophylaxis to those treated also with six-months of antibiotic prophylaxis but with the addition of EF 2) to assess changes in the urinary microbiome between these two groups; and 3) to determine the durability of clinical outcomes in each of the treatment arms up to two years following the six-month intervention (**Figure 2**).

**Figure 1.**
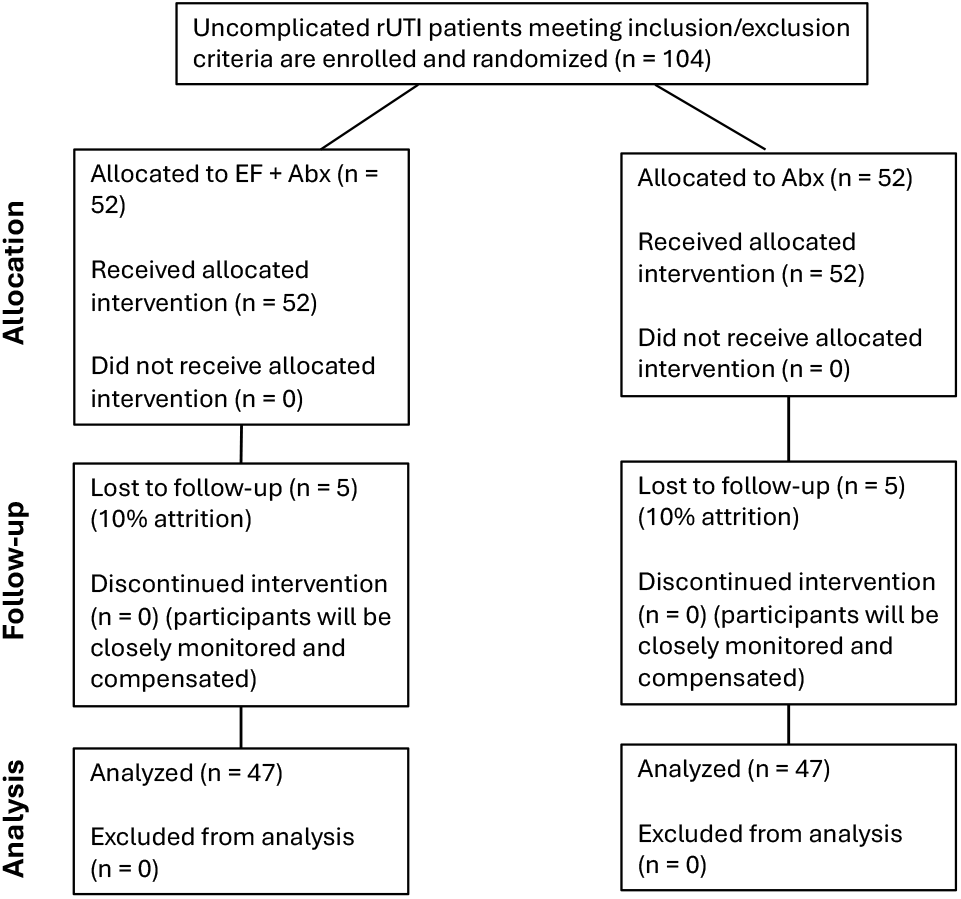
Flow diagram.

**Figure 2.**
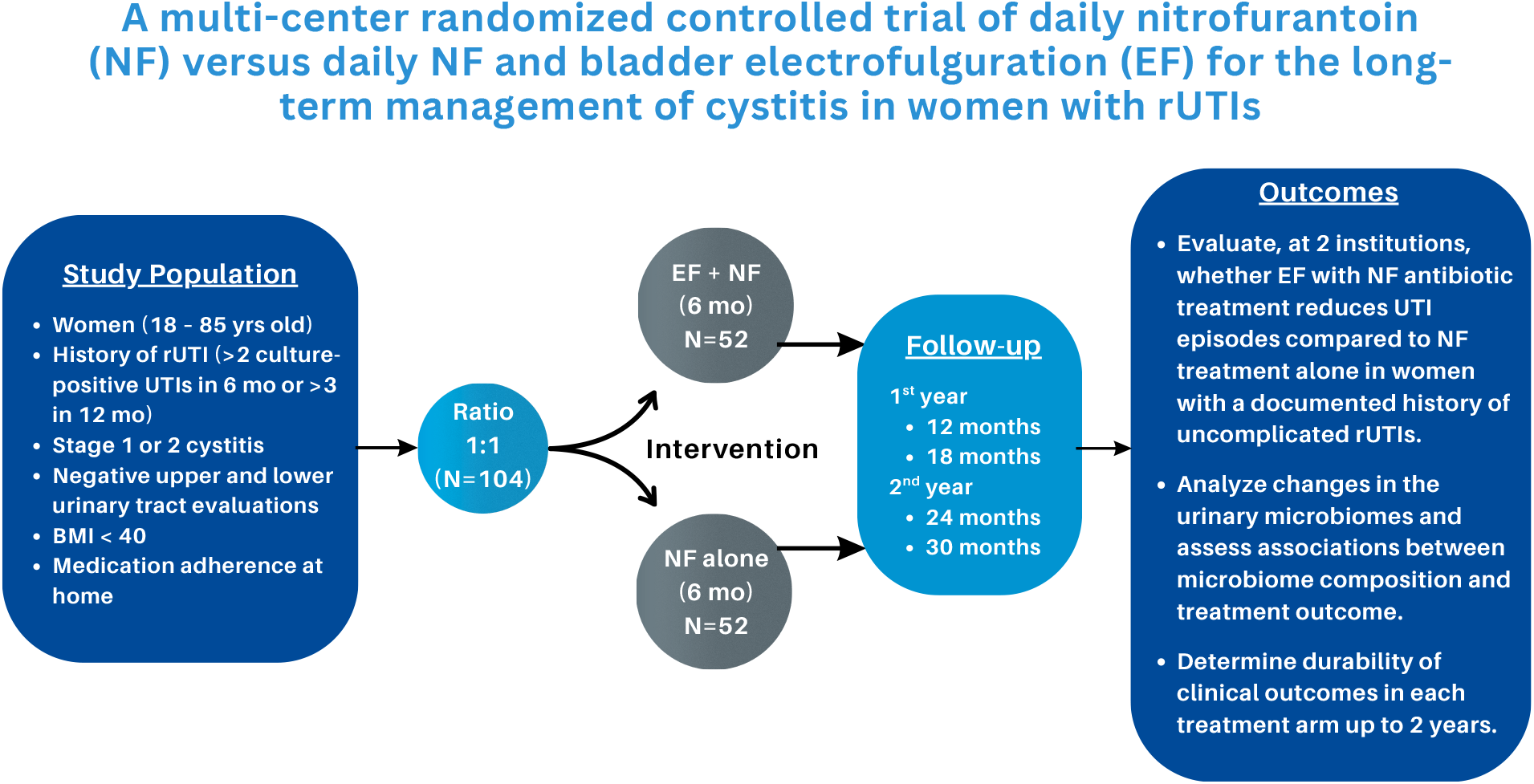
Overall study design methodology.

The trial design and reporting were developed in accordance with the CONSORT recommendations for randomized trials. Participants are randomized to a group using permuted blocks of variable length (2/4/6/8). The randomization scheme has been prepared by a statistician and has been integrated inside a password protected REDCap database shared by both participating centers. Written informed consent is obtained from each woman who enrolls in the study. The trial will take approximately four years (two years of enrollment and two years minimum follow-up) and began enrolling patients in April 2026.

### Antibiotic Treatment

Consistent with current guidelines^14^ on the management of rUTI, the study uses NF at a 100mg dose daily for antibiotic prophylaxis. Participants are directed to take their daily NF antibiotic prophylaxis in the evening to increase compliance,^15^ and NF is administered for six months following EF and for the same six-month duration in the NF alone group.

Nitrofurantoin is a preferred antibiotic for rUTI treatment and prevention due to its narrow spectrum solely indicated to treat UTIs, rapid concentration in the urine, low rates of resistance and allergy, low cost, and favorable safety profile.^16,17^ In our prior study on the impact of antibiotic allergy and resistance in older women with rUTIs, we showed that for nearly one-third of women NF was the only viable treatment option.^6^ In addition, both centers have tested and reported on the comparable efficacy of NF to treat most bacterial strains causing UTIs at their two institutions.^17^

### Electrofulguration Procedure

Electrofulguration^18,19^ is a brief outpatient procedure done with a female cystoscope with a blunt-end sheath and a fine tip bugbee electrode using a very low current to superficially cauterize cystitis lesions. The procedure takes less than 30 minutes on average. Participants void before being discharged and are asked to start their daily low-dose of NF (100mg) immediately following the procedure. Photographs documenting extent of cystitis are obtained before and after EF. **Figures 3** and **4** depict findings from cystoscopy before and after EF treatment and have been published previously.^13,20^ Post-operatively, patients are informed that they may experience increased urgency and occasional bladder spasms for a few days. Other complications such as clot retention or pain are exceedingly rare. Bladder perforation and ureteral injury are exceedingly rare and have not been observed in prior published series.^21^

**Figure 3.**
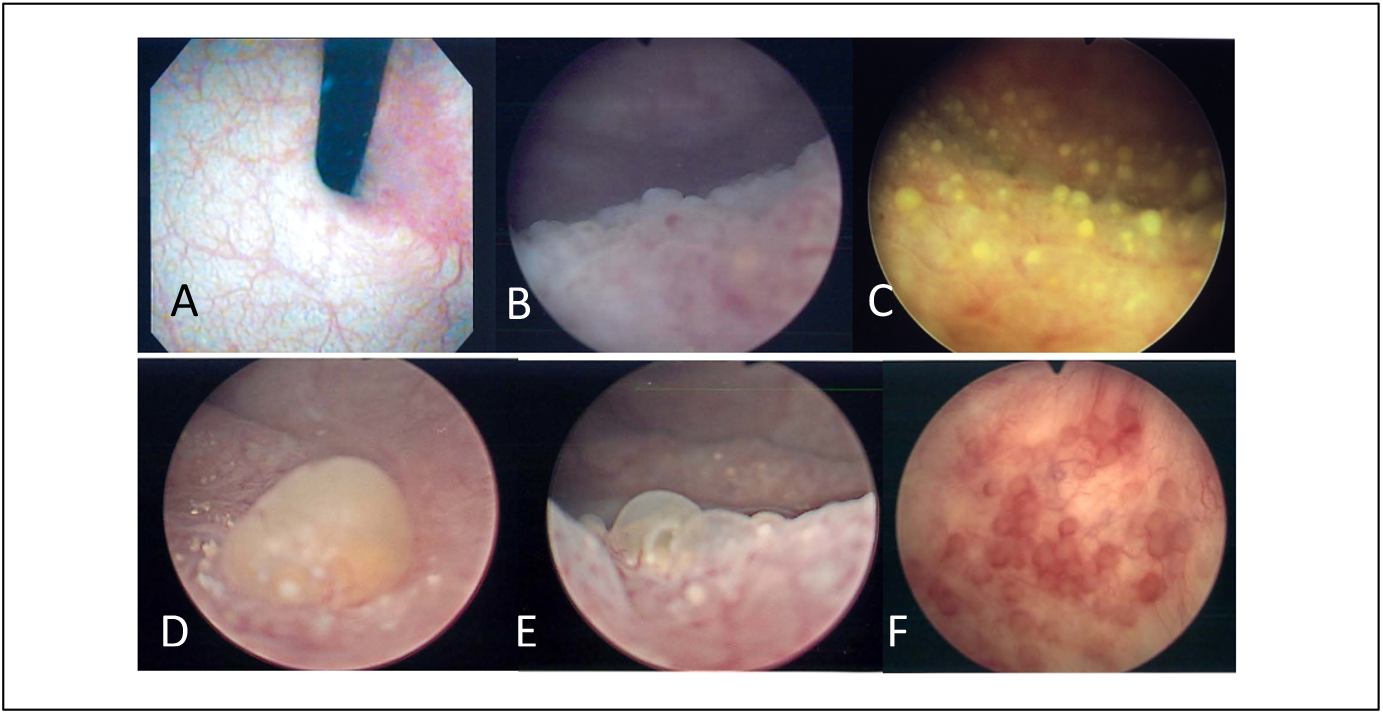
Cystoscopy findings from Ordonez *et al*. PMID 35944653. A) Retroflex view of stage 1 chronic trigonitis. B) Chronic trigonitis stage 1 with cystitis cystica. C) and D) Pus pockets over trigonitis area. E) Stage 2 lesions involving the trigone and bladder base. F) Cystitis cystica of the bladder base (stage 2).

**Figure 4.**
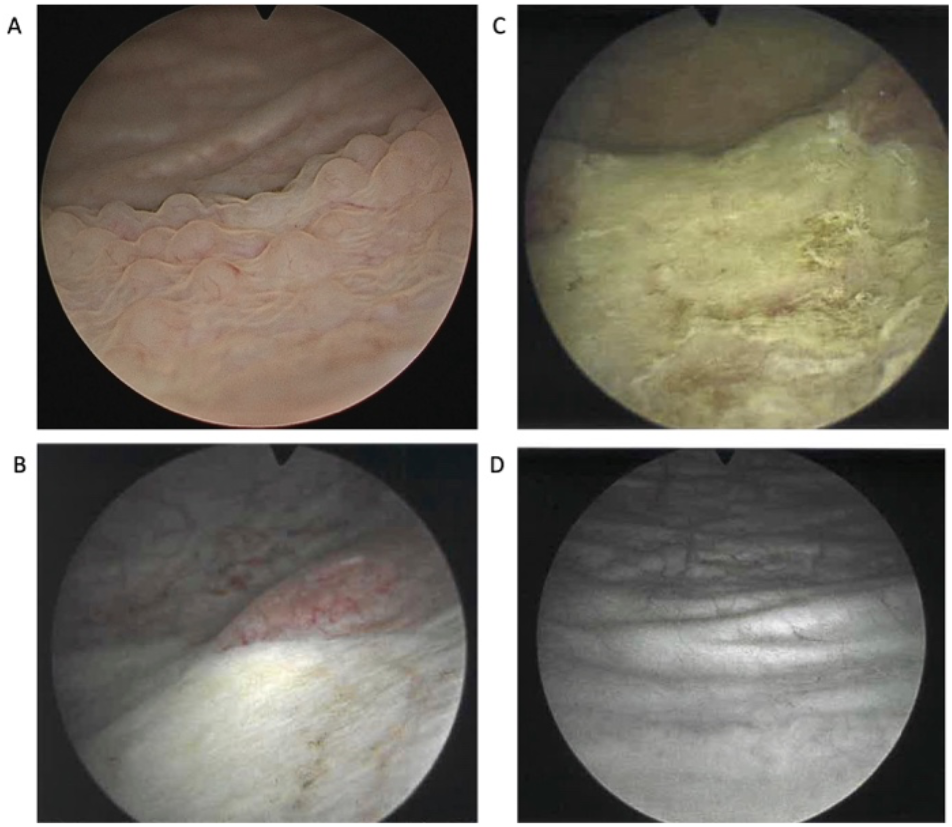
Cystoscopy findings from Gaitonde *et al*. PMID: 37384844. A) Chronic cystitis cystica. B) Left side of the trigone just after fulguration. C) Appearance of the whole trigone from the level of the bladder neck just after fulguration. D) Six months after fulguration—healed trigone and bladder base areas.

### Study Population

Female patients 18 to 85 years old with history of uncomplicated rUTI for at least the past year will be screened for inclusion and exclusion criteria (**Table 1**). Eligible participants provide written informed consent at the baseline visit. Included in the criteria is a negative upper and lower urinary tract evaluation, including pelvic examination for pelvic organ prolapse (less than or equal to stage 2), measurement of post-void residual (less than 50 mL), and imaging (which may include renal ultrasound and standing voiding cystourethrogram) to exclude kidney stone, hydronephrosis, reflux, or urethral diverticulum.

**Table 1.**
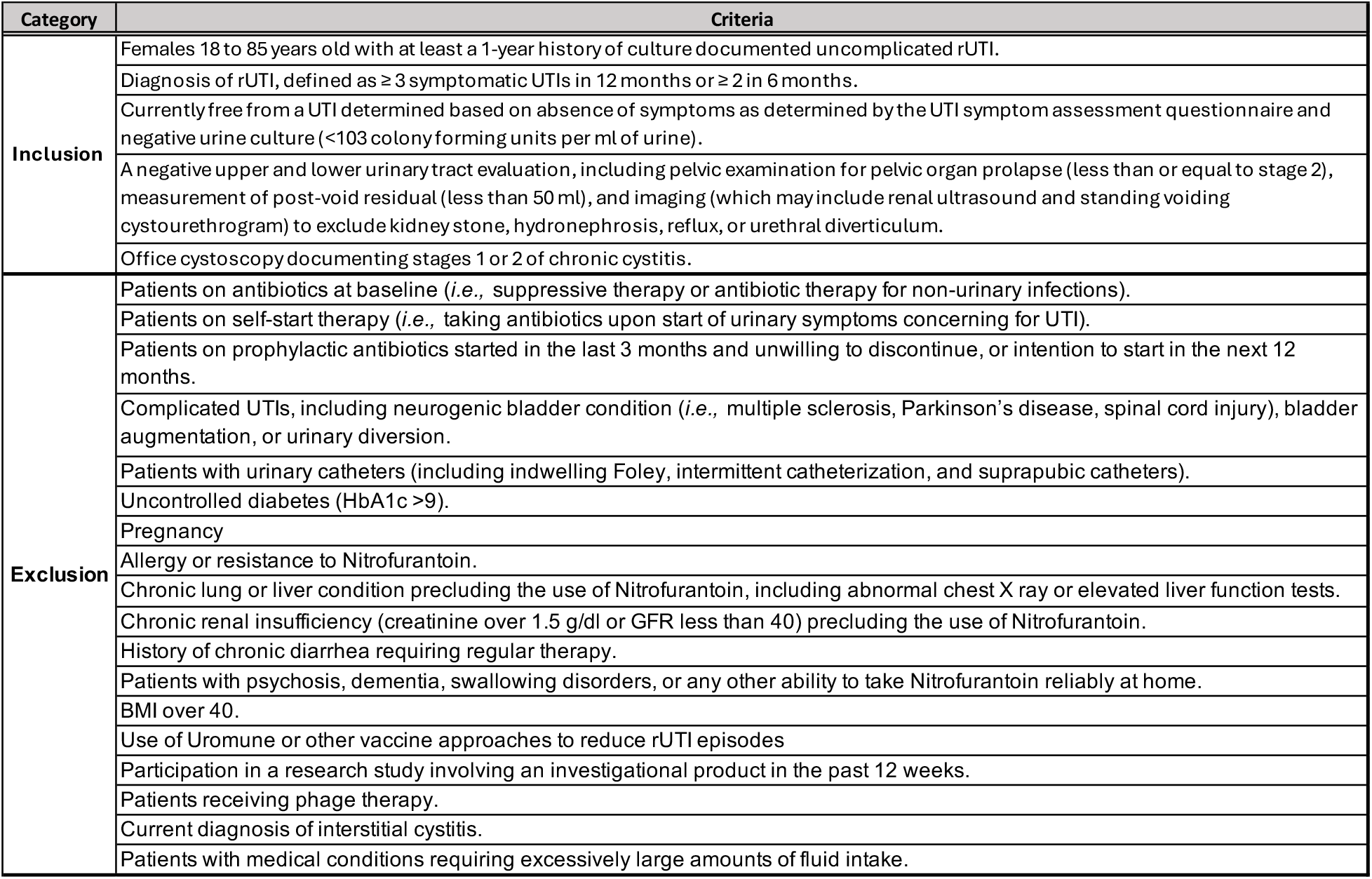
Inclusion and exclusion criteria.

### Primary Outcome

The primary outcome in this trial is the rate of culture-documented symptomatic UTI episodes following the completion of each six-month study arm intervention. Culture-documented UTIs will be tabulated from 6 to 18 months (first year) and 18 to 30 months (second year for measure of durability) after the completion of the original intervention. The urinary microbiome will be analyzed at baseline and over the duration of the study to assess the effect of each intervention on the prevalence of uropathogenic species versus uroprotective species in the urinary tract. Age, urine pH, bacterial species, stage of cystitis (1 versus 2), hormonal status, sexual activity, and diabetic status represent the covariates in this study. Failure to remain UTI-free is determined based on the presence of UTI symptoms documented by validated questionnaires and documented urine culture confirming a bacterial species associated with UTI at each UTI episode following the end of the 6-month intervention arm.

### Secondary Outcomes

A number of secondary outcomes are also being considered. We are comparing patient characteristics between the two enrolling institutions, as well as the rate of culture-documented breakthrough symptomatic UTIs and urine cultures showing multidrug resistant organisms in each treatment arm. The rate of non-urine culture documented symptomatic UTI episodes for 12 months and for 24 months following the completion of treatment arms, as determined by the UTISA^22^ (a total score of 3 or higher, or any individual score of 2 or 3); the rate of behavior changes in both groups (fluid increases or use of non-antibiotic therapies, e.g., cranberry, D-mannose, urinary analgesics, hormonal therapy); and the rate of score differences in patient symptoms and quality of life (QoL) assessments using validated questionnaires— the UTISA,^22^ UDI-6,^23^ and QoL by VAS^24^ will also be evaluated between the treatment arms. Additionally, the rate of UTI episodes (after the first and second year of the completion of the intervention) compared to the rate of UTI episodes in the year prior to the start of the study and the number of patients who have adverse events/side effects (severe defined as those requiring withdrawal of treatment; mild defined as those not requiring withdrawal of treatment) will be tabulated in both groups.

### Assessment Schedule

Urine samples are collected during office visits, which occur at baseline (week 0), 6 weeks, 3 months, 6 months, and at 12 months and 18 months (year 1), and 24 and 30 months (year 2). We collect midstream clean-catch urine from study participants as shown in **Table 2** during the initial office visit and the 6-, 12-, and 18-month visits.

**Table 2.** Overview of study procedures and timeline.

| Action | Month |  |  |  |  |  |  |  |
| --- | --- | --- | --- | --- | --- | --- | --- | --- |
|  | Baseline | 1.5 | 3 | 6 | 12 | 18 | 24 | 30 |
| Informed Consent | X |  |  |  |  |  |  |  |
| Inclusion/Exclusion Criteria | X |  |  |  |  |  |  |  |
| Urine collection for microbiome studies | X |  |  | X | X | X |  |  |
| Urine collection for urinalysis/pH, culture | X | X | X | X | X | X | X | X |
| Cystoscopy and imaging of the trigone | X |  |  | X |  |  |  |  |
| Chest x-ray and liver function test | X |  |  | X |  |  |  |  |
| Receive antibiotic treatment | X | X | X |  |  |  |  |  |
| Patient quality of life assessments using validated questionnaires | X | X |  |  |  | X | X | X |
| Patient symptoms assessments using validated questionnaires | X | X | X | X | X | X | X | X |

#### Baseline visit

Participants in each group provide urine samples for microbiome analysis and undergo baseline chest X-ray and liver function tests (LFTs). Those with abnormal LFTs or chronic lung lesions on chest X-ray at baseline are excluded from the study. If randomized into the EF group, patients undergo cystoscopy under light general anesthesia and fulguration of the trigone (stage 1) or trigone and bladder base (stage 2). Images are taken before and after and participants are directed to start their daily low-dose of NF (100mg) immediately following the procedure.

#### Six-week visit

All participants review NF tolerance, confirm medication compliance with the Morisky scale,^25,26^ provide samples for urinalysis (UA) and urine culture, answer the three questionnaires (UTISA, QoL, UDI-6), and receive a 6-week supply of NF pills. Participants in the EF group are queried regarding their post-procedure recovery.

#### Three-month visit

Participants in both groups will review their NF tolerance, confirm medication compliance with the Morisky scale, provide samples for UA, urine culture, and microbiome assays, answer three questionnaires (UTISA, QoL, UDI-6), and receive a 3-month supply of NF pills.

#### Six-month visit

Participants in both intervention group review NF tolerance, confirm medication compliance (Morisky scale), provide samples for UA, urine culture, and microbiome assays, and answer three questionnaires (UTISA, QoL, UDI-6). All participants undergo repeat chest X-rays and LFTs. All participants undergo office cystoscopy to document changes in areas of chronic cystitis noted at baseline. Cystoscopy studies are video recorded in our EMR and graded by blinded investigators.

#### 12 and 18 months, 24 and 30 months

These visits represent 6- and 12-months from the end of the intervention and 18 and 24 months from the end of the intervention, respectively. Primary and secondary outcome measures will be reported at this time. Study failures are defined as two urine culture-documented symptomatic UTI episodes in the first six months or three UTI episodes in the first year.

### Sample Collection

Urinary creatinine concentration is measured to allow urine concentration normalization. All samples are stored immediately at 4°C. Urine is aseptically aliquoted and stored at -80°C within three hours of collection. Urine is glycerol-stocked for cultivation of the living microbiota for validation of sequencing results and future mechanistic studies and molecular studies.

### Telephone Follow-up

Participants in each group are contacted via telephone between clinic visits. At each time point, they will be queried with validated questionnaires and asked about new episodes of UTI and adverse events.

### Measures

#### Assessment of UTI

The presence of a UTI is based on symptoms documented by validated questionnaires—the Urinary Tract Symptom Assessment (UTISA) questionnaire,^22^ Urinary Distress Inventory short form (UDI-6),^23^ and QoL by Visual Analog Scale (VAS),^24^ and also documented urine culture confirming the bacterial species present at each UTI episode following the end of the 6-month intervention arm. In real life practice, we expect some patients will have UTI symptoms late at night or during the weekend. They may receive an antibiotic treatment for such event depending on the severity of these symptoms. Such non-culture documented events are recorded, but do not have the same meaning for failure calculations as a urine culture-documented episode. To encourage patients to utilize our clinical management during such an event, we provided personal cards with the study team phone numbers to use any time during the study, and encourage each participant to provide a urine sample before initiating a new antibiotic prescription. To that effect, participants received a home urine sample collection kit to store their urine overnight, as well as placing standard urine culture orders at their preferred, close to home, lab facilities. At times, participants may opt for a courier (Eagle Express) to promptly deliver their home urine sample to the lab.

#### Interpretation of urine culture

Diagnosing culture-confirmed symptomatic UTIs among study participants and distinguishing from asymptomatic bacteriuria will be executed by a team of providers who specialize in rUTIs, antimicrobial resistance, and antibiotic stewardship.

#### Study treatments

This study is limited to two treatment groups: NF alone and NF following EF. Each of the two groups receives daily NF for six months. If a participant experiences a breakthrough UTI, they will be placed on culture-directed antibiotics and subsequently, resume their prophylaxis unless their culture indicates NF resistance, in which case their participation is terminated. The study protocol is clearly defined by each visit to ensure standardization between providers and sites.

### Statistical Design

#### Power Analysis and Sample Size Estimation

Power and sample size estimations were calculated using PASS 2024 software. For the EF + NF and NF groups, survival probabilities of 50% (*instead* of 65%, the mean of 60%-70%)^13,27^ and 25% (mean of 20%-30%)^28,29^ were used, respectively, in the analysis. Using a smaller survival probability of 50% for the EF + NF group allows for more uncertainty and results in a more conservative hazard ratio estimate of 0.5 (note: the estimated hazard ratio would have been 0.31 using the higher survival probability of 65% for the EF + NF group). A two-sided log rank test with a type I error rate of 5% requires an overall sample size of 90 subjects (45 in the EF + NF group, 45 in the NF group) to achieve 80% statistical power to detect a hazard ratio of 0.5 when the proportions surviving in the EF + NF and NF groups are 50% and 25%, respectively. The study lasts for 54 months of which subject accrual occurs in the first 24 months. The accrual pattern across time periods is uniform (all periods equal). To account for a 10% dropout rate, an overall sample size of 104 subjects (52 in the EF + NF group and 52 in the NF group) is needed, which increases the power to 85%. The parameters used in the power analysis included: recruitment time (24 months); time-to-follow-up (30 months); total time (54 months); and allocation ratio (1:1).

### Statistical Analyses

Treatment success will be evaluated using Kaplan-Meier analysis and the log rank test to assess survival times of EF + NF and NF groups. As a secondary measure, the Cox proportional-hazards model will be implemented to determine the association between the hazard (as a function of time) of time-to-failure in EF + NF and NF groups while accounting for menopausal status and other characteristic variables.

The least absolute shrinkage and selection operator (LASSO) variable selection method will be used to select the best set of predictors associated with patient survival times in the Cox models. Descriptive statistics for patient characteristics will be presented as medians and interquartile ranges for continuous measures, and frequencies and percentages for categorical measures. For baseline measurements, EF + NF, and NF groups will be compared using the t-test for continuous variables and the chi-square test for categorical variables. If assumptions are not met, then nonparametric alternatives such as the Mann-Whitney U test for continuous variables and Fisher’s exact test for categorical variables will be implemented instead. For repeated measurements, EF with NF and NF alone groups will be compared using linear mixed effects models for continuous outcomes and generalized estimating equations (GEE) for categorical outcomes. Each patient will be treated as a random effect. Additional factors such as menopausal status and all covariates will be evaluated. When a mixed effects model is significant, appropriate post hoc tests are run to identify where differences occur. All analysis assumptions are carefully evaluated, with adjustments made for multiple testing when appropriate. All statistical tests will be two-sided using a type I error rate of 5%. All analyses will be conducted using R software version 2026.01.2+418.

## Discussion

Examining the effects of EF with NF versus NF alone will address the hypothesis that those receiving EF in addition to long-term NF therapy will fare better after one year. Should this result, the clinical trial described herein would demonstrate the added advantage of EF over antibiotic prophylactic management alone. To ensure the generalizability of the findings, the study is being conducted at two institutions. Also, to confirm the durable benefits of EF from existing published reports from our group and others, as well as provide new long-term data on the benefit of NF alone, the study will extend to two years of follow-up for each group.

### Choice of Procedural Intervention

We have shown that bladder lesions harbor tissue-embedded bacterial reservoirs,^12^ and based on extensive studies in mouse models of UTI, we hypothesize that these reservoirs serve as sources of reinfection.^30–32^ Our prior work shows that, in many patients, EF may eliminate tissue-associated bacterial reservoirs that contribute to reinfection.^8–11,13^ Because EF specifically targets the bladder mucosa, we hypothesize that the urinary microbiome composition changes as a result of EF. We further hypothesize that eliminations of tissue-embedded uropathogenic reservoirs within the bladder and the subsequent resolution of cystitis, may result in a transition to a more beneficial urinary microbiota in patients receiving EF.

### Choice of Antibiotic

It is generally well established that long-term antibiotic use can change the composition of the gut and vaginal microbiota.^33,34^ However, the impact of antibiotic therapy on the urinary microbiome is not well characterized.^35^ Further, the impact of antibiotics prescribed for the management of rUTI on the urinary microbiome has not been investigated longitudinally. It is important to monitor the impact of antibiotics prescribed for rUTI on the urinary microbiome because it is a known reservoir for uropathogens, and the presence of a beneficial urinary microbiome (i.e., lactobacilli) is associated with protection against UTI.^36^ This study presents an unprecedented opportunity to study the impact of NF, which is a first-line, and guideline-recommended antibiotic for the management of rUTI, on the urinary microbiome. In addition to its broad mechanism of action, which makes development of resistance more difficult among uropathogens, NF has been demonstrated to reach bactericidal concentrations only in urine and not in plasma, suggesting that it may selectively target urinary or bladder-resident bacteria.^37^ Previous work has only evaluated the impact of NF on cultivable uropathogens (mainly *E. coli*) in vaginal and fecal samples.^1,29^ These data along with the documented pharmacokinetics of NF absorption and excretion lead us to the hypothesis that urinary microbiome composition will be significantly altered during NF therapy. Longitudinal analysis of the urinary microbiome before and after NF treatment will, for the first time, elucidate the long-term effect of NF on the urinary microbiome.

### Impact on Microbiome

The effects of NF alone, or EF followed by NF on urinary microbiome composition has not been previously assessed in humans. Based on previous clinical observations; however, we expect to detect differences in urinary microbiome composition between baseline and after antibiotic treatment for both arms. We predict the following outcomes: microbial communities of the urine may stabilize in both cohorts between the 6- and 12-month time points due to withdrawal of the antibiotics; EF will remove bacterial reservoirs (of presumably uropathogenic bacteria) within the bladder epithelium such that lower relative abundances of uropathogenic taxa (e.g., *E. coli, Klebsiella pneumoniae, E. faecalis*, among others) will be detected in the urine of postmenopausal women; and importantly, we anticipate our mediation analysis to identify potential causal relationships between EF responsive urinary microbiome taxa in each niche and outcomes (positive or negative) of the efficacy of EF in preventing rUTI.^38^ Identification of potentially causal taxa associated with positive or negative outcomes, will be important not only for identifying individuals who may benefit from EF, but will also be critical to the design of prebiotic and probiotic therapies that may boost efficacy of EF.

### Durable Response Effect

The current guideline-recommended management of women with stage 1 and/or 2 cystitis and a history of uncomplicated rUTIs is based on long-term (six months) antibiotic prophylaxis therapy.

Research has evidenced a favorable and durable response to EF treatment in women with stage 1 and stage 2 cystitis and a history of uncomplicated rUTI.^13^ Antibiotic usage decreased following EF, suggesting that UTI incidence may have also declined. Therefore, we hypothesize that EF in combination with antibiotic prophylaxis may confer more durable clinical benefit by targeting complementary pathogenic mechanisms.

### Outcome Measures

The optimal method for assessing outcomes in women with history of uncomplicated rUTI and early stages of cystitis remains unclear. However, monitoring for UTI episodes and using cystoscopy to visualize lesions in the bladder wall remain a viable option for evaluating the effects of an intervention on rUTIs and chronic infection. This study leverages EF as a supplement to the standard of care (long-term antibiotic prophylaxis) for this population.

### Potential Limitations

This study will be conducted at two centers, which could increase the generalizability of our results. The study population may not be generalizable to all patients, however, as many participants may be attending a Urogynecology and Reconstructive Pelvic Surgery (URPS) subspecialty clinic. Further, despite our efforts to recruit a diverse population and translate consent forms in Spanish, the study population might involve more Caucasian, postmenopausal women, which could also limit generalizability. The duration of this study may allow for introduction of confounding variables that may affect participant’s UTI rates, including new onset of medical comorbidities. Socio-environmental factors that arise during the study period and behavior changes could confound results and be difficult to capture. Regular telephone contact with participants will be used to inquire about this type of change, but it is possible that all instances are not being captured. We anticipate these uncaptured changes will occur with the same frequency in each treatment group.

## Conclusion

Recurrent UTIs impose substantial physical, psychological, and economic burdens, compounded by antibiotic resistance and limited durability of standard therapies. Observational work suggests that targeted EF of chronically inflamed bladder regions, quite possibly harboring resident bacteria, could reduce recurrence of UTI episodes. This randomized clinical trial will offer level I evidence as to whether EF in combination with NF antibiotic prophylaxis is superior to standard of care and guideline-recommended prophylaxis with NF alone in reducing UTI episodes with a 2-year follow-up after the end of the intervention period. Moreover, the study will enable longitudinal data on alterations of the urinary microbiome across treatment modalities and assessment of its association with patient outcomes.

## Data Availability

All data produced in the present study are available upon reasonable request to the authors

## Acknowledgements

The authors would like to acknowledge Dr. Sarah Mason Eck, Ph.D., for the preparation and review of this manuscript.

